# Wearable Personal PM_2.5_ Monitoring During Wildfire Smoke Episodes in Metro Detroit

**DOI:** 10.64898/2026.07.30.26359253

**Authors:** Clara G. Zundel, Thomas Fikes, Erika Strobel, Megan Schrimpf, Hilary Marusak

**Author notes:** **Corresponding Author:** Clara G. Zundel, 3113-577-4520, 3901 Chrysler Service Dr., 5^th^ floor, Room 571, Detroit, MI 48201.

## Abstract

Wildfire smoke has increasingly affected air quality across North America, raising concerns about the health effects of fine particulate matter (PM_2.5_) exposure, including potential impacts on brain health. However, relatively few studies have characterized personal PM_2.5_ exposure during these events using wearable monitoring. We examined daily personal PM_2.5_ concentrations during wildfire smoke episodes in southeast Michigan alongside neighborhood outdoor PM_2.5_ estimates. Four participants (one adolescent and three adults) wore AirBeam3 personal monitors during ongoing studies. Neighborhood outdoor PM_2.5_ was estimated using the average of three nearest PurpleAir outdoor air quality sensors, and wildfire smoke days were identified using state air quality advisories. Group-level descriptive statistics summarized personal and neighborhood outdoor PM_2.5_ and self-reported time spent outdoors. Exploratory within-participant analyses quantified associations between neighborhood outdoor and personal PM_2.5_ concentrations on smoke and non-smoke days. Neighborhood outdoor daily PM_2.5_ concentrations were higher during wildfire smoke days than non-smoke days (87.6 ± 80.2 vs. 12.3 ± 7.0 µg/m^3^). Personal PM_2.5_ concentrations were more than five times higher during wildfire smoke days (28.2 ± 20.0 vs. 5.1 ± 4.7 µg/m^3^) than non-smoke days. Within participants, every 10 µg/m^3^ increase in neighborhood PM_2.5_ was associated with 2.3 µg/m^3^ increase in personal PM_2.5_ concentrations. Wearable PM_2.5_ monitoring captured elevated personal exposures while providing individual-level exposure information beyond neighborhood outdoor air quality estimates. These findings demonstrate that wearable monitoring complements neighborhood air quality measurements by capturing individual-level exposure, providing a more comprehensive assessment of real-world wildfire smoke exposure for future studies examining the effects on brain health.

**Societal Significance Statement:** Wildfire smoke is becoming more frequent and severe, exposing millions of people to unhealthy levels of fine particulate matter (PM_2.5_). Most public air quality information comes from outdoor monitoring stations, but these measurements do not necessarily reflect the amount of pollution an individual breathes. In this study, personal PM_2.5_ concentrations were generally lower than neighborhood outdoor air quality estimates, yet they increased more than fivefold during wildfire smoke events, even though participants reported spending relatively little time outdoors. Overall, more than 25% of participant-days exceeded the United States Environmental Protection Agency (EPA) 24-hour PM_2.5_ standard. These findings suggest that reducing time outdoors may lessen, but not completely eliminate, wildfire smoke exposure. As evidence continues to emerge linking wildfire smoke exposure to respiratory, cardiovascular, and brain health, combining wearable personal air pollution monitors with neighborhood air quality measurements may help researchers and public health officials better understand individual exposure, support the evaluation of protective strategies such as indoor air filtration and cleaner-air spaces, and inform recommendations that better protect communities during future wildfire smoke events.

## Introduction

Wildfires are increasing in frequency, duration, and intensity across North America, contributing to more frequent episodes of poor air quality in communities located both near and far from active fires^1^. Fine particulate matter (PM_2.5_), a major component of wildfire smoke, can be transported hundreds to thousands of kilometers, resulting in episodic smoke events that affect populations well beyond the fire source^2^. In addition to well-established respiratory and cardiovascular effects, growing evidence suggests that PM_2.5_ exposure may influence brain health through mechanisms including neuroinflammation and oxidative stress, with potential implications for cognitive and mental health outcomes^3^.

Despite growing concern regarding the health impact of wildfire smoke, accurately characterizing individual PM_2.5_ exposure remains challenging. Most epidemiologic studies estimate wildfire smoke exposure using fixed-site regulatory monitoring networks, satellite-derived products, or low-cost ambient sensor networks, such as PurpleAir^4^. While these approaches provide valuable information on regional and neighborhood air quality, they may not accurately reflect the PM_2.5_ concentrations individuals experience throughout their daily activities. Personal exposure is influenced by factors including time spent indoors and outdoors, building infiltration, ventilation, and individual activity patterns, resulting in substantial variability even among people living in the same community^5^. Wearable personal air quality monitors offer an opportunity to directly characterize individual PM_2.5_ exposure during real-world wildfire smoke events by capturing exposures across daily activities and microenvironments.

Several recent studies have reported associations between wildfire smoke exposure and adverse neurobehavioral outcomes, including reduced attention and cognitive performance in adults, as well as altered patterns of frontoparietal activity during cognitive processing^6,7^. A recent review further highlights growing evidence linking wildfire smoke to cognitive, neurological, and mental health outcomes while identifying important gaps in understanding exposure-response relationships^4^. However, most studies to date have relied on wildfire disaster exposure classification (i.e., direct, indirect, or witnessed exposure) or ambient air quality estimates rather than objective measures of *personal* PM_2.5_ exposure, limiting the ability to characterize the exposures individuals experience. Improved characterization of individual PM_2.5_ exposure during wildfire smoke events is an important step toward refining estimates of health risk and supporting future investigations into the neurobehavioral consequences of wildfire smoke exposure.

Therefore, the objective of this study was to characterize personal PM_2.5_ exposure measured using wearable personal monitors during two documented wildfire smoke events in southeast Michigan and describe these measurements alongside neighborhood outdoor PurpleAir estimates. By directly measuring personal PM_2.5_ exposure during naturally occurring wildfire smoke episodes, this study provides real-world insight into the variability in individual exposure under typical daily living conditions.

## Methods

### Study Design

This descriptive secondary analysis used personal PM_2.5_ monitoring data collected during two independent environmental health studies conducted in the Detroit metropolitan area. Neither parent study was designed to investigate wildfire smoke exposure. However, personal monitoring periods from a subset of participants coincided with two documented Canadian wildfire smoke events affecting southeast Michigan, providing an opportunity to characterize personal PM_2.5_ exposure during these episodic events. All study procedures were approved by the Wayne State University Institutional Review Board, and participants (or parents/legal guardians for minors) provided written informed consent prior to participation. Minors provided assent.

### Participants

Participants were drawn from two independent environmental health studies conducted in the Detroit metropolitan area. Study 1 recruited adolescents aged 10-17 years between 2023 and 2024 to examine associations between environmental PM_2.5_ exposure and emotional health. Study 2 recruited adults aged 19-69 beginning in 2025 and ongoing to investigate associations between environmental exposures and cardiovascular health. Participants from both studies completed seven consecutive days of personal PM_2.5_ monitoring using wearable AirBeam3 monitors. Monitoring periods that overlapped documented Canadian wildfire smoke events (June 7-8, 2023, and July 15-20, 2026) were included in the present analysis. Participant characteristics, monitoring dates, and valid monitoring days are presented in **Table 1**.

**Table 1.**
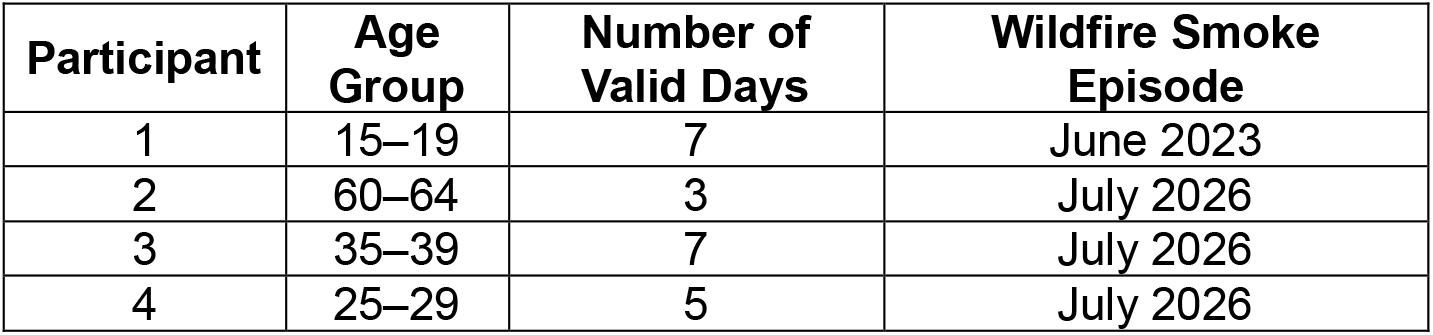
Characteristics of participants included in the descriptive analysis of personal PM_2.5_ exposure during documented wildfire smoke events.

| Participant | Age Group | Number of Valid Days | Wildfire Smoke Episode |
| --- | --- | --- | --- |
| 1 | 15–19 | 7 | June 2023 |
| 2 | 60–64 | 3 | July 2026 |
| 3 | 35–39 | 7 | July 2026 |
| 4 | 25–29 | 5 | July 2026 |

### Personal PM_2.5_ Monitoring

Personal PM_2.5_ exposure was measured using the AirBeam3 (HabitatMap, Brooklyn, NY), a portable wearable air quality monitor equipped with a Plantower PMS7003 optical particle sensor. The AirBeam3 has demonstrated good agreement with Federal Equivalent Method (FEM) reference instruments in independent laboratory and field evaluations^8^. Participants were instructed to wear the monitor throughout their daily activities for seven consecutive days, removing it only during activities that could damage the device (e.g., bathing or swimming) and while sleeping. Monitors operated in standalone mode, recording second-by-second PM_2.5_ concentrations (µg/m^3^) and corresponding GPS location data directly to an onboard microSD card. Daily mean personal PM_2.5_ concentrations were calculated from all valid monitoring data for each monitoring day.

Daily PM_2.5_ summaries were calculated only for days meeting the wear-time criterion, defined as at least 1,080 minutes (75% of 24-hour period) of valid AirBeam3 measurements. Days not meeting the wear-time criterion were excluded from analysis.

### Neighborhood Outdoor PM_2.5_ Assessment

Neighborhood outdoor PM_2.5_ concentrations were estimated using data from the PurpleAir sensor network (PurpleAir Inc., Draper UT), which provides continuous ambient PM_2.5_ measurements from low-cost optical particle sensors. For each participant, the three outdoor PurpleAir sensors nearest to their residence were identified and used throughout the monitoring period. Daily mean PM_2.5_ concentrations (PM2.5_cf_1) and corresponding relative humidity measurements were downloaded using the PurpleAir Data Download Tool.

Daily mean PM_2.5_ concentrations were averaged across the three sensors, and the U.S. Environmental Protection Agency (EPA) correction equation^9^ was subsequently applied using the corresponding daily mean relative humidity values to estimate neighborhood outdoor PM_2.5_ concentrations.

### Identification of Wildfire Smoke Episodes

Wildfire smoke episodes were identified *a priori* using official Air Quality Alerts issued by the Michigan Department of Environment, Great Lakes, and Energy (EGLE) for southeast Michigan and corresponding public health advisories from the Detroit Health Department attributing degraded air quality to smoke transported from Canadian wildfires^10–13^. Participant monitoring days were classified as wildfire smoke days if they overlapped official air quality alerts and public health advisories issued in response to Canadian wildfire smoke. This included the June 7-8, 2023, EGLE Air Quality Action Days for elevated fine particulate matter in southeast Michigan^11,12^, or the multiple statewide EGLE Air Quality Alerts issued between July 15-20, 2026^10,13^.

### Daily Surveys

Participants completed a brief daily survey each evening during the seven-day monitoring period to capture contextual information related to their daily activities. For the present analyses, only the self-reported time spent outdoors was used to provide context for personal PM_2.5_ exposure. Participants reported the approximate amount of time they spent outdoors each day using one of seven response categories: 0 minutes, <10 minutes, 10-30 minutes, 31-59 minutes, 1-2 hours, 2-3 hours, >3 hours.

### Statistical Analysis

All analyses were conducted in R (version 4.6.1). Descriptive statistics were used to summarize participant characteristics monitoring compliance, and daily personal and neighborhood outdoor PM_2.5_ concentrations. Daily mean PM_2.5_ concentrations were summarized separately for wildfire smoke and non-smoke days using means, standard deviations, and ranges, as appropriate. Time spent outdoors was also summarized descriptively to provide context for personal exposure patterns.

As an exploratory analysis, linear regression models were used to estimate within-participant associations between (1) wildfire smoke day (smoke vs. non-smoke) and daily personal PM_2.5_ concentrations and (2) neighborhood outdoor PM_2.5_ concentrations and daily personal PM_2.5_ concentrations. Participant ID was included as a categorical covariate in each model to account for differences in baseline PM_2.5_ concentrations between participants, thereby estimating associations using within-participant comparison.

## Results

### Participant monitoring and wildfire smoke episodes

Four participants (one adolescent and three adults) completed monitoring during wildfire smoke episodes identified through official state air quality alerts. Across the four participants, 28 participant-days of monitoring were attempted. Twenty-two participant-days (78.6%) met the wear-time criterion and were included in the analysis, including 13 wildfire smoke days and 9 non-smoke days.

### Personal PM_2.5_ during documented wildfire smoke episodes Group-level descriptive findings

At the group level, neighborhood outdoor PM_2.5_ concentrations were substantially higher during wildfire smoke days than on non-smoke days (M: 87.6 ± 80.2 vs. 12.3 + 7.0 µg/m^3^; **Table 2**). Personal PM_2.5_ concentrations were more than five times higher during wildfire smoke days (M: 28.2 ± 20.0 vs. 5.1 ± 4.7 µg/m^3^; **Table 2**; **Figures 1-2**).

**Figure 1.**
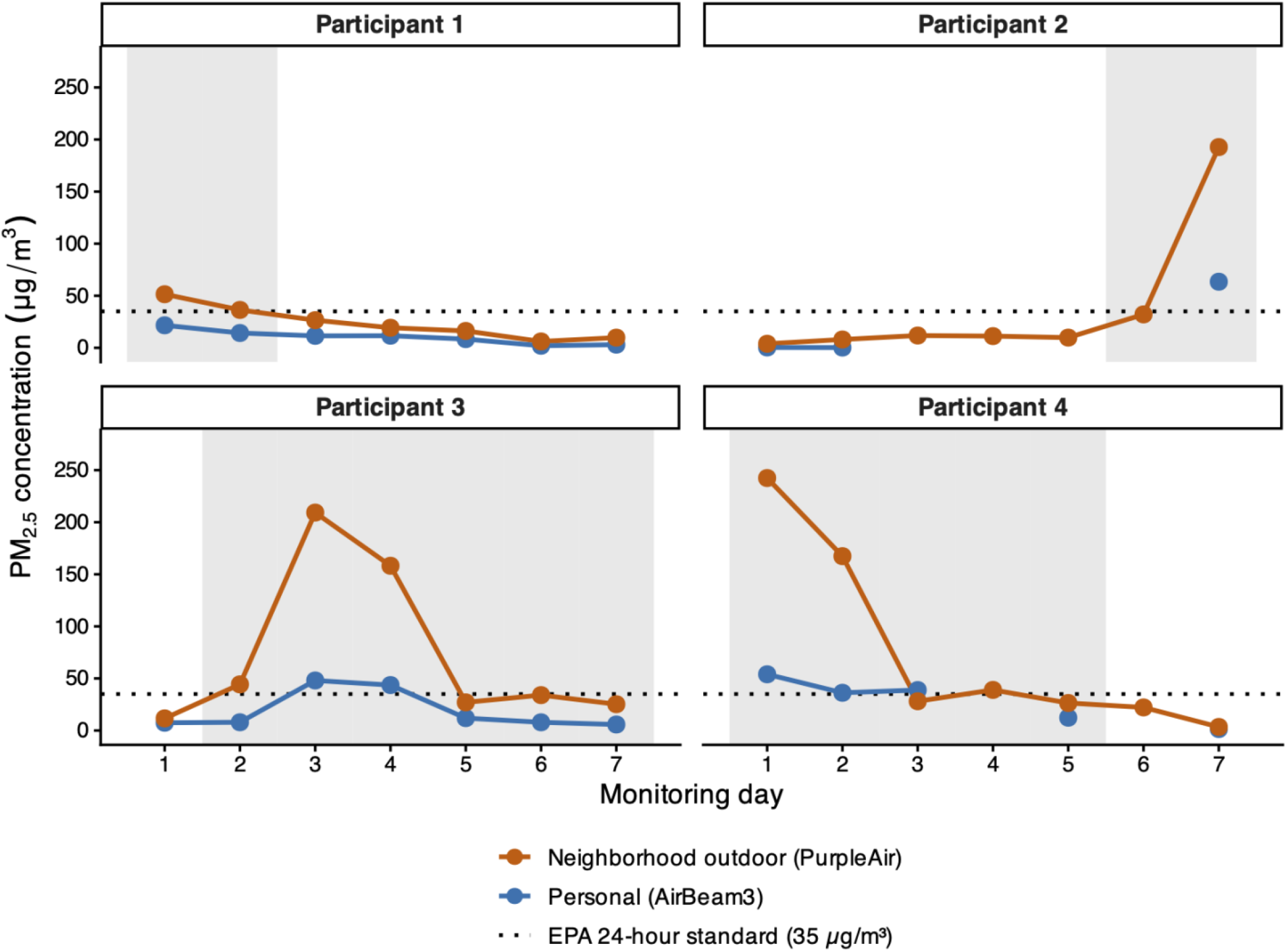
Daily mean personal PM_2.5_ concentrations measured using wearable AirBeam3 monitors and neighborhood outdoor PM_2.5_ concentrations estimated from the average of the three nearest PurpleAir outdoor sensors. Gray shading denotes days classified as wildfire smoke days based on official state air quality alerts. Each panel represents one participant. The horizontal dotted line indicates the U.S. Environmental Protection Agency (EPA) 24-hour National Ambient Air Quality Standard (NAAQS) for PM_2.5_ (35 µg/m^3^) and is shown for reference. Missing values represent days that did not meet the minimum AirBeam 3 wear-time requirement for inclusion in the analysis.

**Figure 2.**
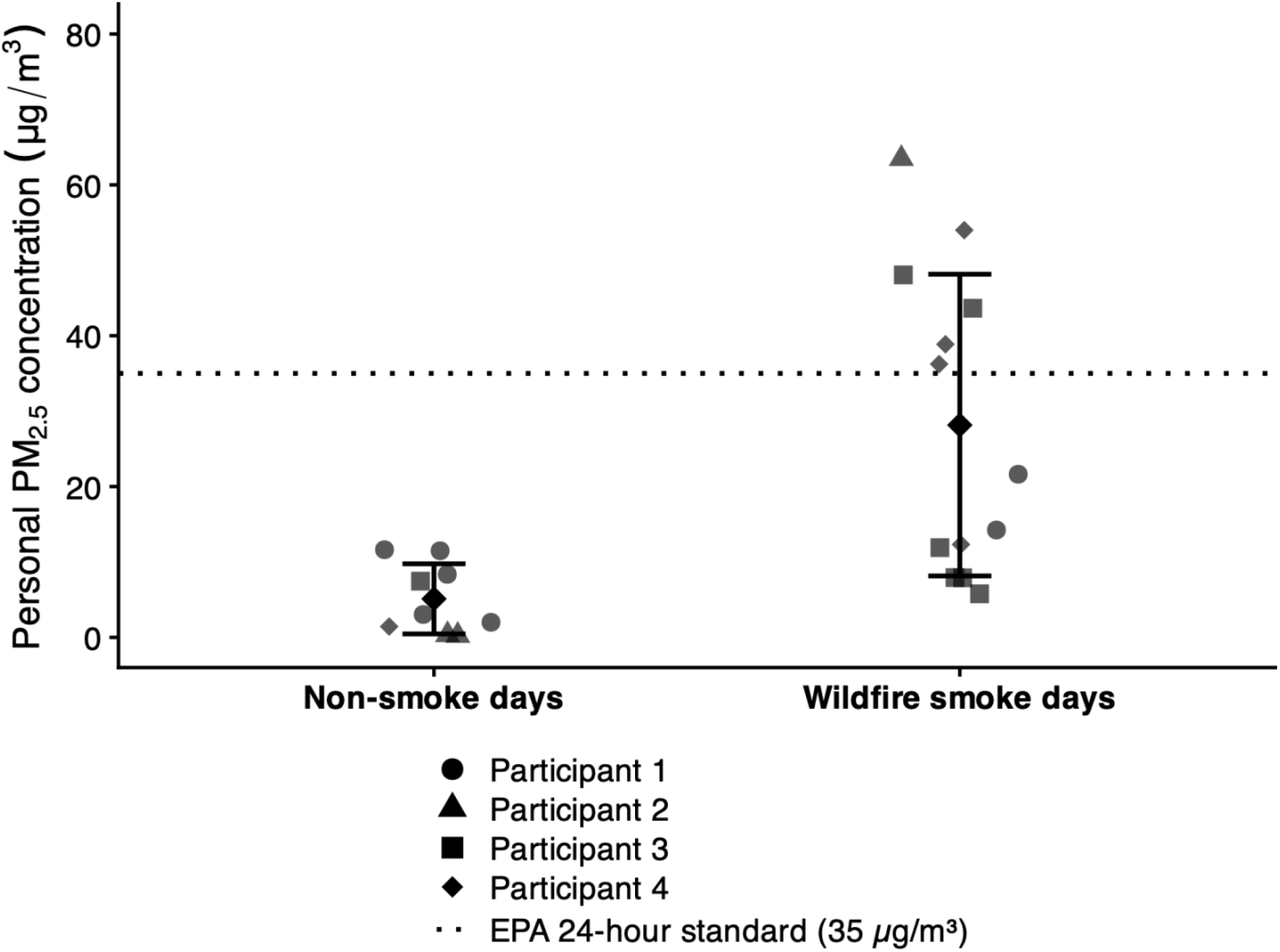
Daily mean personal PM_2.5_ concentrations measured using wearable AirBeam3 monitors during non-smoke and wildfire smoke days. Individual points represent valid participant-days, with point shapes denoting individual participants. Black diamonds indicate the mean, and error bars represent + 1 standard deviation. The horizontal dotted line indicates the U.S. Environmental Protection Agency (EPA) 24-hour National Ambient Air Quality Standard (NAAQS) for PM_2.5_ (35 µg/m^3^) and is shown for reference.

**Table 2.** Daily neighborhood outdoor and personal PM_2.5_ concentrations and self-reported time spent outdoors during wildfire smoke and non-smoke monitoring days.

| Characteristic | Non-smoke days | Wildfire smoke days |
| --- | --- | --- |
| Participant-days, n | 9 | 13 |
| Neighborhood outdoor PM<sub>2.5</sub> (µg/m<sup>3</sup>), mean ± SD | 12.3 ± 6.98 | 87.6 ± 80.2 |
| Personal PM<sub>2.5</sub> (µg/m<sup>3</sup>), mean ± SD | 5.1 ± 4.7 | 28.2 ± 20.0 |
| Personal PM<sub>2.5</sub> range (µg/m<sup>3</sup>), mean ± SD | 0.2 – 11.6 | 5.8 – 63.5 |
| Self-reported time outdoors |  |  |
| 0 mins | 0 (0.0%) | 0 (0.0%) |
| <10 mins | 3 (33.3%) | 7 (53.8%) |
| 10-30 mins | 4 (44.4%) | 1 (7.7%) |
| 31-59 mins | 0 (0.0%) | 2 (15.4%) |
| 1-2 hours | 0 (0.0%) | 2 (15.4%) |
| 2-3 hours | 0 (0.0%) | 0 (0.0%) |
| >3 hours | 2 (22.2%) | 1 (7.7%) |

Daily personal PM_2.5_ concentrations during wildfire smoke days ranged from 5.8 to 63.5 µg/m^3^, and personal PM_2.5_ concentrations exceeded the U.S. Environmental Protection Agency (EPA) 24-hour PM_2.5_ National Ambient Air Quality Standard of 35 µg/m^3^ on 27.3% of participant-days (**Figures 1-2**). In contrast, personal PM_2.5_ concentrations remained comparatively low and exhibited less day-to-day variability on non-smoke days.

Participants generally reported spending less time outdoors during wildfire smoke days than during non-smoke days (**Table 2**). More than half (53.8%) of wildfire smoke monitoring days involved less than 10 minutes spent outdoors, whereas longer outdoor durations, including more than 3 hours outdoors, were reported more frequently on non-smoke days (22.2% vs. 7.7%).

### Exploratory within-participant analyses

Within participants, each 10 µg/m^3^ increase in neighborhood outdoor PM_2.5_ was associated with a 2.3 µg/m^3^ increase in personal PM_2.5_ concentrations (B = 2.3, 95% CI [1.7, 2.8], *p* < 0.001). Within participants, daily personal PM_2.5_ concentrations were, on average 25.6 µg/m^3^ higher on wildfire smoke days than on non-smoke days (B = 25.6, 95% CI [9.1, 43.0], *p* = 0.007).

## Discussion

In this study, we demonstrated the utility of using low-cost wearable air pollution monitors to characterize personal PM_2.5_ exposure during real-world wildfire smoke episodes. Personal PM_2.5_ concentrations were more than 5 times higher during wildfire smoke days than non-smoke days which tracked with higher neighborhood outdoor PM_2.5_ concentrations. Consistent with this finding, within participants, every 10 µg/m^3^ increase in neighborhood outdoor PM_2.5_ was associated with a 2.3 µg/m^3^ increase in personal PM_2.5_ concentrations. Although personal PM_2.5_ concentrations generally remained lower than neighborhood outdoor estimates, 27.3% of participant-days exceeded the U.S. EPA 24-hour PM_2.5_ National Ambient Air Quality Standard, despite participants reporting relatively little time spent outdoors. The lower personal concentrations relative to neighborhood outdoor estimates may reflect the fact that individuals spent substantial portions of the day indoors, where building infiltration and air filtration can reduce exposure to outdoor-derived PM_2.5_, although these factors were not directly measured in the present study. Together, these findings highlight that neighborhood outdoor PM_2.5_ concentrations alone do not fully characterize personal exposure during wildfire smoke events, supporting the complementary use of wearable monitoring in environmental health research.

These findings are noteworthy given growing evidence that PM_2.5_ exposure may affect not only cardiopulmonary health, but also systemic inflammation and mental health. Growing evidence identifies PM_2.5_ as an important environmental determinant of brain health across the lifespan, with associations reported for neurodevelopmental outcomes during childhood as well as cognitive decline and neurodegenerative disease in older adults^14–17^. Accurate characterization of individual-level PM_2.5_ exposure is therefore critical for understanding exposure-health relationships and identifying populations at greatest risk, particularly as climate-driven wildfire smoke events become increasingly frequent. Consistent with this growing body of literature, prior studies conducted by our group demonstrated that ambient PM_2.5_ concentrations well below the EPA 24-hour PM_2.5_ standard of 35 µg/m^3^ were associated with altered functional neural connectivity within and between major attention networks, greater anxiety symptoms, and elevated inflammatory markers among adolescents^18,19^. Although these studies assessed different exposure periods and cannot be directly compared with the daily personal measurements reported here, they suggest that neurobehavioral and inflammatory changes may occur at PM_2.5_ concentrations substantially lower than those experienced during some wildfire smoke days.

The considerable variability in personal exposure also suggests that outdoor air quality alone does not fully characterize an individual’s exposure. Time spent outdoors, infiltration of smoke into the home, building ventilation, filtration, transportation, and other indoor sources may all influence personal PM_2.5_ concentrations^5^. Public health advisories commonly recommend remaining indoors and limiting outdoor activities during wildfire smoke events. While these behaviors may reduce exposure for many individuals, several participant-days in the present study exceeded the EPA 24-hour PM_2.5_ standard despite participants reporting minimal time spent outdoors. This variability underscores the need to better understand how housing characteristics, ventilation, filtration, and individual behaviors influence personal exposure, particularly as wildfire smoke increasingly affects regions of the United States that have not historically experienced frequent smoke events.

Understanding these exposure modifiers will be important for developing guidance that goes beyond reducing outdoor activity and includes actionable strategies such as improving filtration, creating cleaner-air spaces, and addressing housing-related barriers to smoke protection. Low-cost wearable monitors provide a scalable and accessible approach for characterizing individual-level exposure during wildfire smoke events, complementing fixed-site monitoring networks by capturing substantial heterogeneity in personal exposure. As such, these tools may help evaluate the effectiveness of public health recommendations and identify opportunities to further reduce individual exposure.

Several limitations should be considered when interpreting these findings. First, this secondary analysis included a small convenience sample of four participants, limiting the generalizability of the findings and restricting the robustness of the exploratory statistical analyses. Second, although personal PM_2.5_ exposure was measured directly using wearable monitors, additional factors that likely contribute to exposure variability – including housing characteristics, building ventilation, combustion sources, air filtration, transportation, and other time-activity patterns – were not systematically assessed. Third, time spent outdoors was self-reported using broad categorical response options and may not fully capture individual behaviors that influence personal exposure. Finally, these findings were derived from two wildfire smoke episodes in southeast Michigan and may not generalize to wildfire events of differing intensity, duration, or geographic location. Future studies in larger and more diverse populations that integrate wearable monitoring with detailed assessments of indoor environments and individual behaviors will be important for identifying the primary drivers of personal PM_2.5_ exposure and protecting brain and overall health during wildfire smoke events.

As climate change increases the frequency, duration, and geographic extent of wildfire smoke episodes, improved characterization of individual-level PM_2.5_ exposure using wearable personal monitoring alongside neighborhood outdoor air quality measurements will become increasingly important for refining public health recommendations and understanding the effects of wildfire smoke on brain health across the lifespan.

## Data Availability

The data that support the findings of this study are available from the corresponding author upon reasonable request.

